# Effective subthalamic and pallidal deep brain stimulation – are we modulating the same network?

**DOI:** 10.1101/2021.02.02.21250817

**Authors:** Leon Sobesky, Lukas Goede, Vincent J.J. Odekerken, Qiang Wang, Ningfei Li, Bassam Al-Fatly, Martin Reich, Jens Volkmann, Rob M.A. de Bie, Andrea A. Kühn, Andreas Horn

## Abstract

The subthalamic nucleus and internal pallidum are main target sites for deep brain stimulation in Parkinson’s disease. Multiple trials that investigated subthalamic versus pallidal stimulation were unable to settle on a definitive optimal target between the two. One reason could be that the effect is mediated via a common network. To test this hypothesis, we calculated connectivity profiles seeding from deep brain stimulation electrodes in 94 patients that underwent subthalamic treatment and 28 patients with pallidal treatment based on a normative connectome atlas calculated from 1,000 healthy subjects. In each cohort, we calculated connectivity profiles that were associated with optimal clinical improvements. The two maps showed striking similarity and were able to cross-predict outcomes in the respective other cohort (R = 0.38 at p < 0.001 & R = 0.35 at p = 0.027). Next, we calculated an agreement map which retained regions common of both target sites. Crucially, this map was able to explain an additional amount of variance in clinical improvements of either cohort when compared to the maps calculated on the two cohorts alone. Finally, we tested profiles and predictive utility of connectivity maps calculated from different motor symptom subscores with a specific focus on bradykinesia and rigidity. While our study is based on retrospective data and indirect connectivity metrics, it delivers empirical data to support the hypothesis of a largely overlapping network associated with effective deep brain stimulation in Parkinson’s disease irrespective of the specific target.

## Introduction

Deep brain stimulation (DBS) is an effective treatment option in patients suffering from Parkinson’s disease and two surgical target structures have been established to be most effective.^1,2^ The subthalamic nucleus (STN) and the internal pallidum (GPi) have been reported to result in comparable improvements of motor symptoms in prospective double-blinded trials.^3,4^

A recent study has investigated optimal connectivity profiles for treatment of Parkinson’s disease based on STN-DBS.^5^ The study investigated network profiles of active stimulation sites and their relationships to clinical improvements as measured by the motor part of the Unified Parkinson’s Disease Rating Scale (UPDRS-III). Applying normative connectome data to brain stimulation sites in 95 patients from two centers, the study concluded that effective STN-DBS electrodes would modulate a region within the STN that was functionally connected to large portions of the prefrontal cortex, including supplementary motor area (SMA) and inferior frontal cortex (IFC) and functionally anticorrelated with primary motor cortex (M1). Similar studies accumulated evidence that neuromodulation of a specific network may result in clinical responses specific to a certain symptom (for a review see Horn and Fox^6^). Following this concept of *circuitopathies* – networks with impact on specific symptoms or behaviors – it might be possible to modulate the same network at different stimulation sites to reach similar changes in clinical or behavioral outcomes.^7,8^ As mentioned above, since the 2000s, attempts including large randomized clinical trials have been carried out with the aim of establishing a gold standard – either targeting STN or GPi to treat Parkinson’s disease. However, most clinical studies reached the conclusion that neither of the two can be disregarded as a potential target.^3,4,9,10^ This does not imply that modulating either target would lead to identical effects. For instance, while STN-DBS has led to greater amount of L-Dopa reduction, GPi-DBS was associated with a lower rate of neuropsychiatric side effects.^11^ Still, a natural hypothesis could be that both STN- and GPi-DBS might modulate an *overlapping* functional network in patients suffering from Parkinson’s disease. Needless to say, we know this is the case for the STN and GPi. Both nuclei are crucial integrator hubs in the subcortex and direct interactions between the well-studied indirect and hyperdirect pathways have been established.^12^ We have known at least since the seminal work of Oskar and Cecile Vogt in the 1920ies that GPi and STN are tightly interlinked, forming part of what they referred to as the *striatal system*.^13^ Both nuclei play crucial parts to integrate information within the motor control and action-selection system supported by the basal ganglia.^14^

Here, we use the aforementioned network calculated from a multi-center cohort of 95 STN-DBS patients to cross-predict clinical improvements in a cohort of 28 patients that underwent GPi-DBS.^3,5^ In a next step, we calculate an optimal treatment network from the GPi-DBS cohort and use it to cross-predict outcomes in the original STN-DBS cohort. We compare optimal network profiles of both cohorts and describe commonalities and differences of both maps. Specifically, this is done by ways of a novel method to identify the *agreement network* between both cohorts. In this way, we are able to identify regions that are predictive for positive clinical outcome in both cohorts.

## Materials and Methods

### Patients and Cohorts

In this study, patients with Parkinson’s disease that underwent DBS were retrospectively enrolled. In addition to data from a previously published study (95 STN patients stimulated in two German centers^5^; Berlin and Würzburg), a second cohort of 28 patients that underwent GPi-DBS surgery (subcohort of patients operated at the Amsterdam Medical Center enrolled in a prior clinical study) was included.^3^ For one of the 95 STN-patients, subscore improvement scores were not available, hence the patient was excluded. All patients received brain imaging before and after surgery in form of preoperative MRI and postoperative CT or MRI scans (see table 1 for imaging choice and basic patient demographics). Both target groups where clinically evaluated before and >12 months after surgery, under active stimulation, using the motor features of the Unified Parkinson Disease Rating Scale III (motor UPDRS), after withdrawal from dopaminergic treatment for >12 hours (Med OFF).

**Table 1:** Patient demographics.

| Cohort | Age, yr | Included No.<br>(female) | UPDRS-III<br>Baseline<br>OFF | UPDRS-III<br>Baseline<br>ON | Improve<br>ment % | Improve<br>ment<br>Absolute | Postop<br>Imaging<br>MR/CT |
| --- | --- | --- | --- | --- | --- | --- | --- |
| **GPi**<br>**Odekerken**<br>**2013** | 60($\pm 6$) | 28(10) | 43($\pm 14$) | 34($\pm 12$) | 13($\pm 42$) | 9($\pm 16$) | 0/28 |
| **STN**<br>**Horn 2017** | 60($\pm 8$) | 94(29) | 44($\pm 14$) | 22($\pm 10$) | 47($\pm 23$) | 22($\pm 13$) | 35/59 |

As secondary outcome, we calculated improvements for rigidity (item 22), bradykinesia (items 23-26) and tremor (items 20-21) subscores. Total improvements as well as bradykinesia and rigidity improvements were calculated as %-improvements from baseline. For tremor, the absolute difference was calculated due to a lower baseline in items 20-21 and patients with a baseline score below 2 points were excluded from this subanalysis. Demographic features are displayed in table 1, detailed information about the two cohorts have been published elsewhere.^3,5^

### Localisation and VTA calculation

Electrodes in both cohorts were localized using the Lead-DBS software^15^; www.lead-dbs.org) following the updated pipeline of the second version.^16^ Briefly, preoperative MRI and postoperative CT or MRI scans where linearly co-registered using advanced normalization tools (Advanced Normalization Tools; ANTs, http://stnava.github.io/ANTs/). In order to minimise bias introduced by a nonlinear deformation of the brain due to pneumocephalus, the brain shift correction step in Lead-DBS was carried out.^16^ Multispectral preoperative volumes were then used to compute a spatial normalization warpfield into ICBM 2009b Non-linear Asymmetric (“MNI”) space using the SyN Diffeomorphic Mapping approach implemented in ANTs.^17^ As shown in a recent study, the precision of this normalization protocol may lead to results comparable with manual expert segmentations of the STN and GPi and the method was top-performer in two comparative studies for normalizations of the subcortex.^18,19^ Subsequently, DBS electrodes were localized using the PaCER algorithm for CT volumes or the TRAC/CORE method for MRI volumes.^15,20^ Results were carefully inspected and manually refined, if necessary, using Lead-DBS. Anatomical segmentations of subcortical structures shown in the present manuscript were defined by the DISTAL Atlas using the Lead Group Analysis tool.^21,22^ Electric fields (E-fields) were calculated applying a Finite Element Method (FEM)-based model in each patient.^16^

### Statistical analysis

#### DBS network mapping

Seeding from voxels contained in the E-Field model, a functional connectivity profile was calculated using data from 1,000 healthy subjects acquired within the Brain Genomics Superstruct Project.^23,24^ Values in the E-Fields served as weights to generate the connectivity profile using the *Lead Connectome Mapper* tool included within Lead-DBS. This led to connectivity *fingerprints* for each patient describing (average) positive and negative functional connectivity between the pair of E-Fields and other voxels of the brain (figure 1 A). Clinical improvements were then correlated with connectivity fingerprints in a voxel-wise fashion across the two cohorts following the approach reported in a coherent study published previously.^5^ This resulted in *R-map* models of “optimal” connectivity (figure 1 C). These R-maps denote positive (Pearson’s correlation) coefficients for regions that were positively connected predominantly to electrodes of top responders and negative coefficients for the ones predominantly connected to electrodes in poor responding patients.

**Figure 1:**
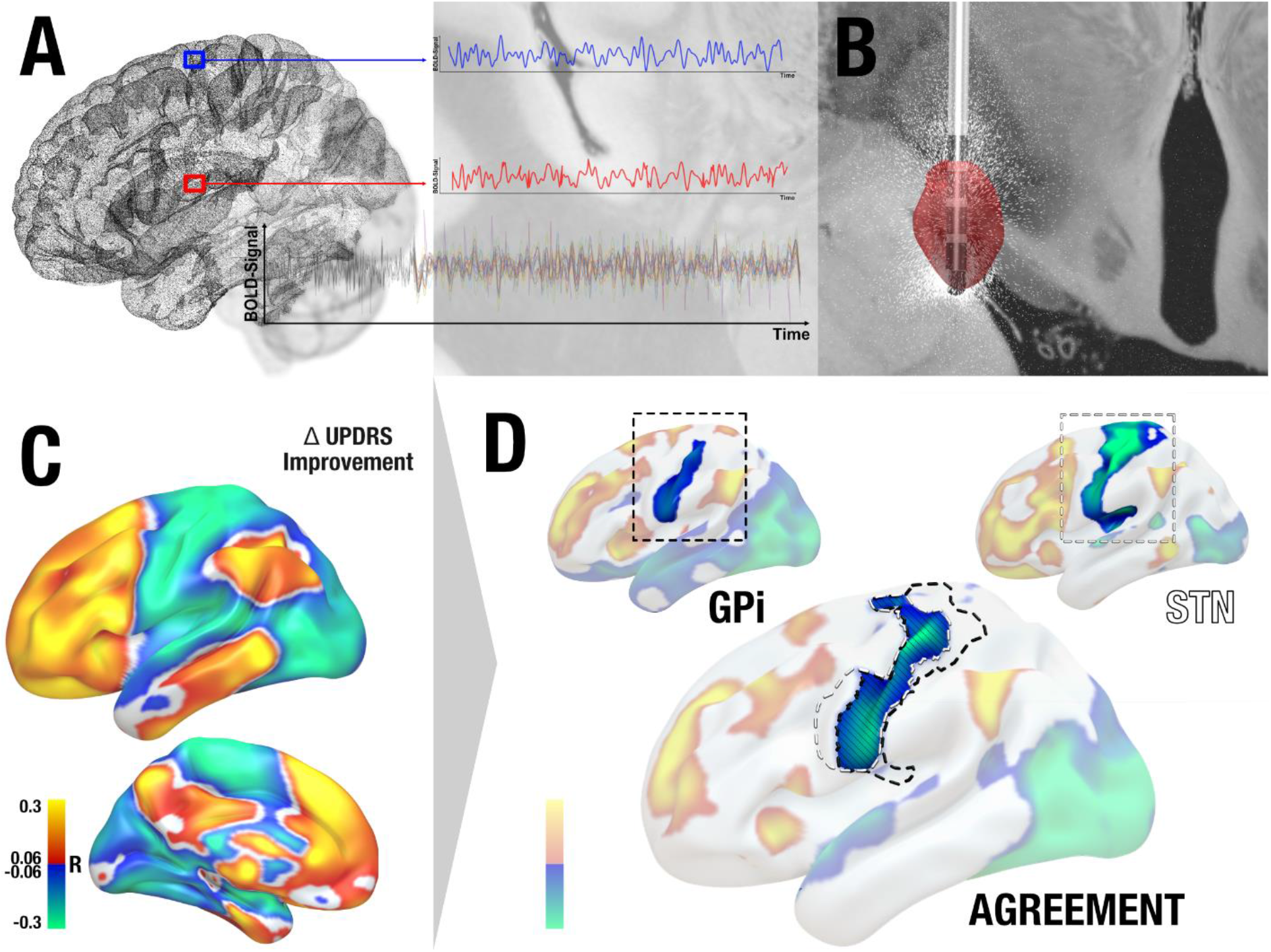
Methods Overview. (A) The connectivity profile of each patient’s VTA is determined by using a functional normative connectome to estimate functional connectivity seeding from the VTA (B) to other areas of the brain. C) This led to connectivity fingerprints for each patient based on which voxel-wise correlation maps with %-improvement scores along the UPDRS were calculated (R-maps). Here, areas to which connectivity was associated with symptom relief are shown in warm colours and the ones associated with poor improvements in cool colours. D) Based on two R-maps, an agreement map was calculated based by retaining areas that had the same sign on both maps (and multiplying their absolute values) and discarding areas in which signs on the two maps were conflicting. This is illustrated in a single example focusing on negative associations with the primary motor cortex.

The R-map model was then used to predict outcomes about clinical improvements in out-of-sample patients. To do so, the connectivity fingerprints (seeding from E-Fields in a specific patient) were spatially compared to the R-map model. The more similar each patient’s fingerprint would be to the R-map, the higher our prediction about their clinical improvements. Here, similarity was calculated by means of spatial correlation across grey matter voxels in the cortex and cerebellum (subcortical voxels were not compared to exclude confounding local effects introduced by electrode placement). Crucially, this prediction step was done in out-of-sample data (in a leave-one-out fashion) to avoid circularity of our prediction model. For instance, the R-map model was calculated on STN-DBS patients to predict outcomes in the GPi sample, and vice versa.

#### Toward an agreement model across DBS targets

Our aim was to show differences and similarities of responsive network profiles in STN- and GPi-DBS. To calculate the set of regions predictive for clinical outcomes *regardless of target choice*, a novel type of map was calculated that we termed *agreement map*. To do so, the R-maps of the two cohorts were superimposed and areas that were either positive or negative in *both* maps were retained (other areas were discarded). To preserve weighting, the remaining values were multiplied across maps while preserving the sign. For instance, if a voxel had values of R = 0.3 and R = 0.2 in the two (STN and GPi) maps, the resulting value in the agreement map would be 0.06. A combination of −0.3 and −0.2 would lead to −0.06 while one of −0.3 and 0.2 would lead to exclusion of the voxel. We hypothesized that regions retained in the agreement map could be more specific to predict clinical outcomes in both cohorts than a model that had seen all data without segregation into the two targets – or a model that was informed by either cohort alone. Hence, we calculated a “conventional” R-map across all datapoints (patients from both the STN and GPi cohort) and compared its predictive utility with the agreement map.

### Data availability statement

The DBS MRI datasets generated during and analyzed during the current study are not publicly available due to data privacy regulations of patient data but are available from the corresponding author upon reasonable request. All code used to analyze the dataset is openly available within Lead-DBS/-Connectome software (https://github.com/leaddbs/leaddbs).

## Results

The STN cohort included 94 patients of 2 independent datasets (29 female, mean age = 60 ± 8 years, with an UPDRS baseline of 44 ± 14 and UPDRS improvement after DBS of 47 ± 23%). The GPi cohort included 28 patients (10 female, mean age = 60 ± 6 years, with an UPDRS baseline of 43 ± 14 and UPDRS improvement after DBS of 13 ± 42%). See Table 1 for further demographic and imaging data. Electrode locations showed DBS electrodes of all patients placed within respective the target regions (figure 2).

**Figure 2:**
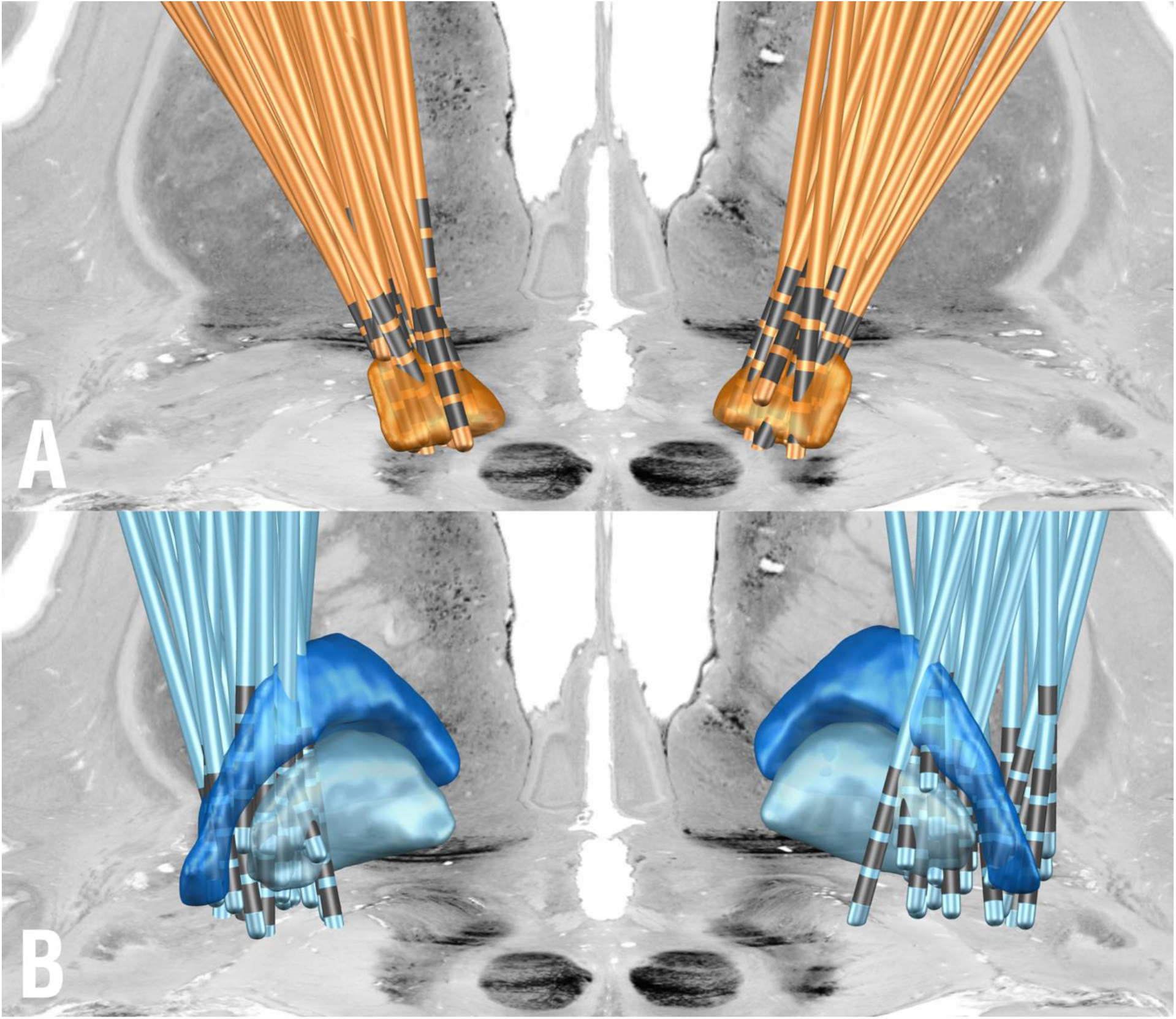
Electrode localizations of the two cohorts shown from posterior. A: STN-cohort described by Horn et al.^5^ B: GPi cohort described by Odekerken et al.^3^ STN: orange, GPi: light blue, GPe: dark blue. Colors of the two cohorts match colors in subsequent figures of the manuscript. In the back, a coronal slice from the BigBrain atlas is shown.^25^

The first R-map model was calculated exclusively on data from the 94 STN-DBS patients (figure 3 top left). Similarity estimates between this map and each whole-brain connectivity fingerprint (seeding from the two DBS electrodes) from the GPi-DBS cohort was calculated and correlated with empirical improvement values in this cohort (figure 3 top right; R = 0.35 at p = 0.027). When repeated vice versa (calculating the R-map model in the GPi sample; figure 3 bottom right; to cross-predict outcomes in the STN sample; bottom left), the correlation was of the same magnitude (R = 0.38 at p = 0.001). In both R-maps, functional connectivity to regions in the frontal lobe, in particular cingulate gyrus, middle and inferior temporal gyri, inferior parietal gyri and motor cerebellum were associated with optimal clinical response (table 2).

**Table 2:** Overlap between spatial correlations and significant connectivity measures. Peak coordinates of STN, GPi and their agreement map (AGR)

| POSITIVE<br>CORRELATIONS |  |  | NEGATIVE<br>CORRELATIONS |  |  |
| --- | --- | --- | --- | --- | --- |
| region | *x/y/z of*<br>GPi<br>STN<br>AGR | t | region | *x/y/z of*<br>GPi<br>STN<br>AGR | t |
| **right**<br>**anterior**<br>**cingulate**<br>**cortex** | 8/32/16<br>16/30/34<br>12/30/30 | 0.53<br>0.53<br>0.16 | **left**<br>**temporal**<br>**lobe** | -42/-30/-16<br>-50/-12/-2<br>-50/-70/-14 | -0.54<br>-0.48<br>-0.14 |
| **left**<br>**superior**<br>**frontal**<br>**gyrus** | -26/56/28<br>-18/4/58<br>-28/56/28 | 0.56<br>0.52<br>0.13 | **left**<br>**precentral**<br>**gyrus** | -40/-4/64<br>-30/-20/52<br>-40/-6/64 | -0.19<br>-0.46<br>-0.05 |
| **left**<br>**cerebellum** | -50/-62/-38<br>-26/-36/-30<br>-50/-62/-38 | 0.43<br>0.49<br>0.1 | **left**<br>**middle**<br>**occipital**<br>**gyrus** | -36/-92/26<br>-4/-98/16<br>-46/-72/10 | -0.42<br>-0.38<br>-0.12 |
| **left**<br>**parental**<br>**lobe** | -50/-52/46<br>-30/-54/40<br>-48/-54/50 | 0.44<br>0.39<br>0.1 |  |  |  |

**Figure 3:**
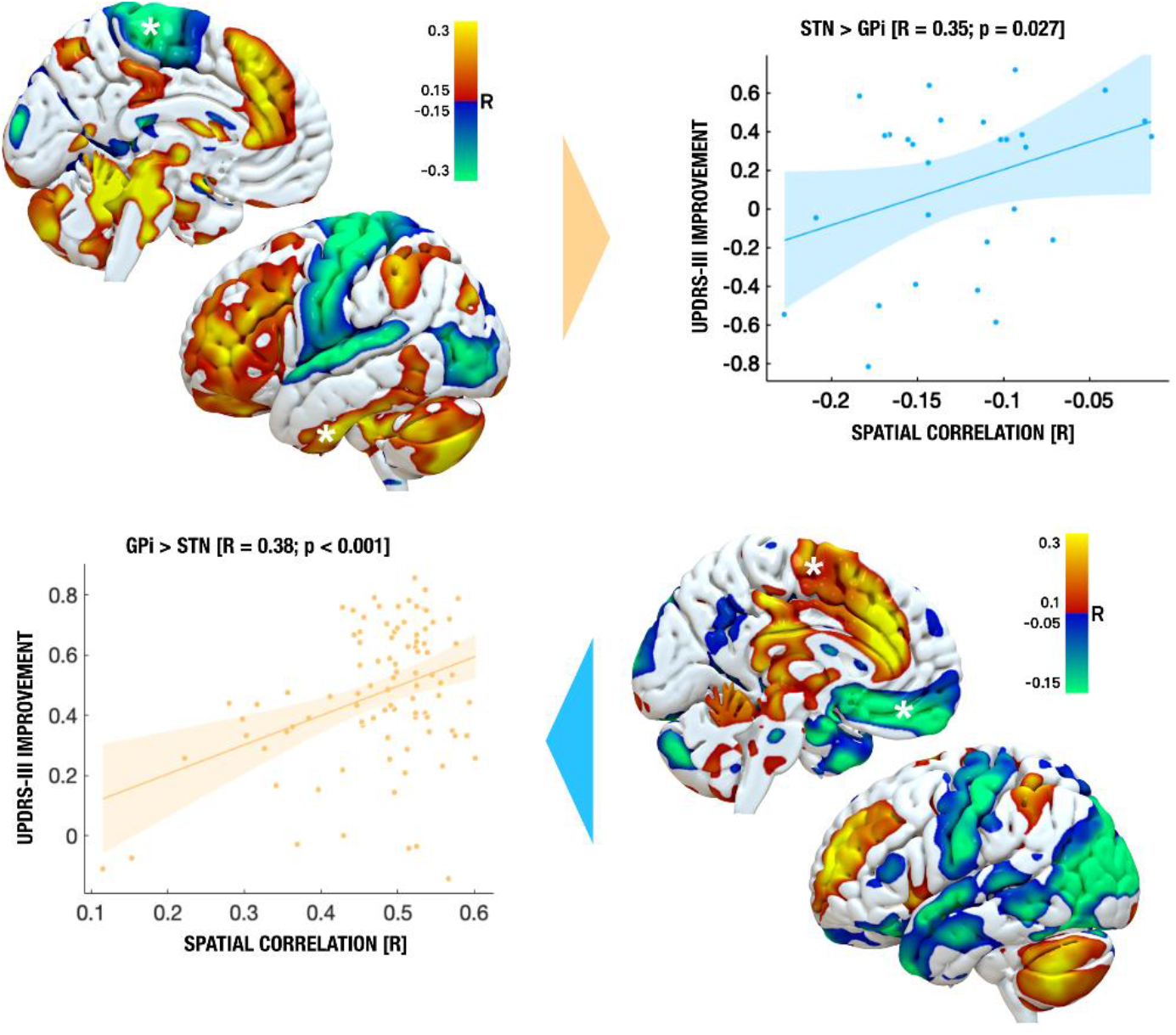
Cross-Predicting DBS outcomes across cohorts and targets. R-maps for both STN-DBS (top left) and GPi-DBS (bottom right) were used to cross-predict clinical outcomes in the other cohort, respectively (rop right shows predictions of outcomes in the GPi-DBS cohort based on the STN-DBS R-map model, bottom left shows the opposite).

In a next step, we aimed at creating a model that would be maximally predictive of clinical outcomes *regardless of* DBS target (STN vs. GPi). A first attempt was to calculate an R-map across the whole group of patients, i.e., simply by correlating clinical outcomes with all 122 patient’s connectivity fingerprints. Not surprisingly, this map (not shown) was again highly similar to the ones shown in figure 3. When using this map to explain variance in clinical outcomes, it was mildly predictive in the STN cohort (R = 0.25 at p = 0.015) but estimates did not significantly correlate with clinical outcomes of the GPi cohort (R = 0.28 at p = 0.15). A subsequent, more deliberate approach was to create an *agreement map* from the two R-maps that were obtained in each cohort, separately (STN R-map and GPi R-map) by discarding regions that did not agree in sign between the two maps (see methods). This type of map is novel in the present context and could intuitively be perceived as a *common denominator* network across the two targets. It was able to significantly explain variance in clinical outcomes in both cohorts (R = 0.37 at p < 0.001 for the STN cohort and R = 0.42 at p = 0.017 for the GPi cohort). Crucially, the amount of variance explained by the agreement map was larger to the one explained by each of the target specific maps (STN and GPi R-map, see figure 4). Hence, by refining the predictive model from data in a second target (albeit with an independent cohort), the amount of explained variance could be increased. Of note, these values are meant for direct head-to-head comparisons and should be interpreted with care since the analysis setup is somewhat circular. However, by integrating these results with leave-one-out predictions (figure 3) and the regions within the agreement map (figure 4), the agreement map could inform a hypothesis of a common treatment network for Parkinson’s disease that will be made openly available to empower further validation in subsequent studies.

**Figure 4:**
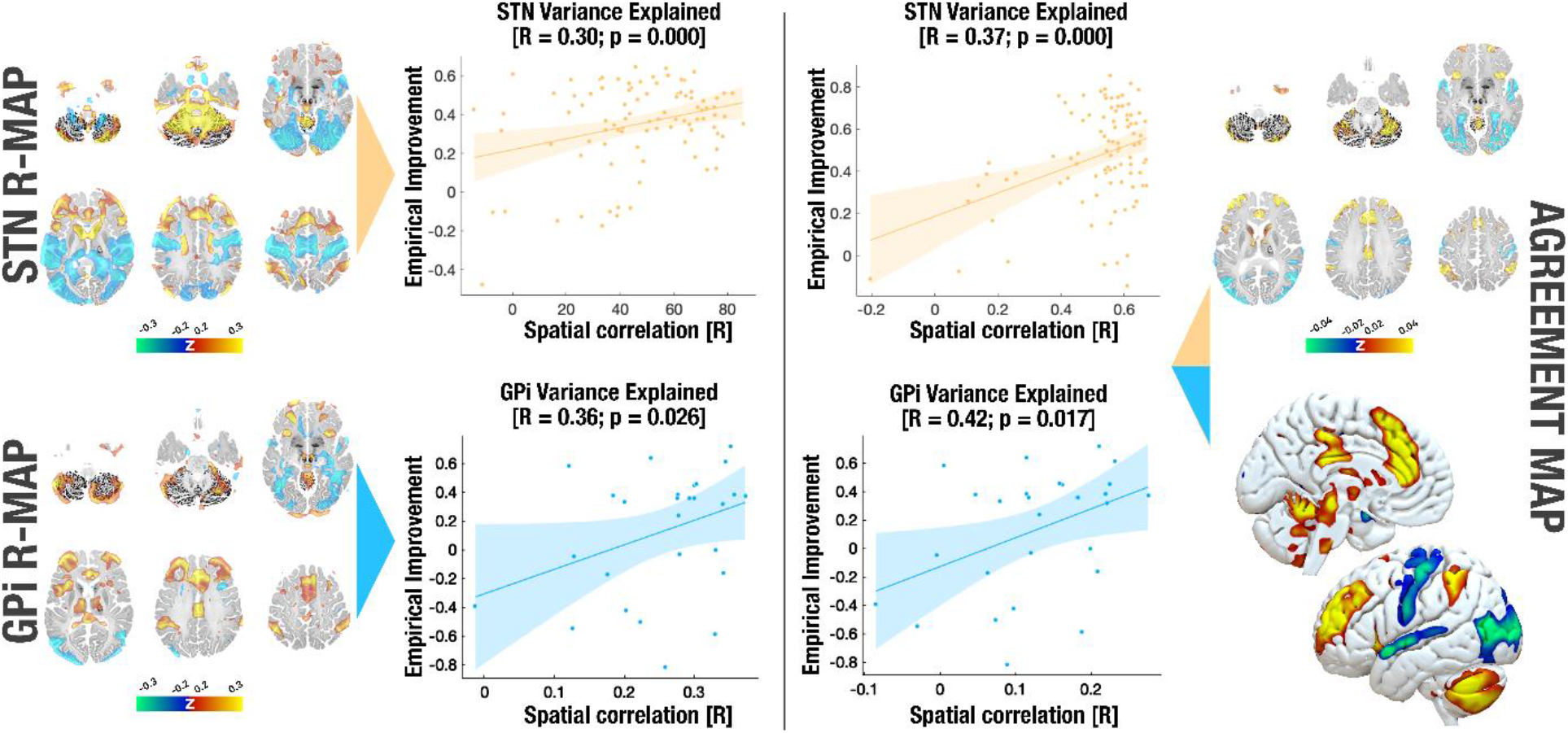
Variance in clinical outcomes explained by different models. Predictive utility of the R-map calculated in each target (left) and an agreement map, in which only regions predictive for both targets were retained (right). Using the the R-map calculated on either target alone, a significant portion of variance in outcomes in both cohorts could be explained (R = 0.30 at p < 0.001 for STN; R = 0.36 at p = 0.026 for GPi). The agreement map was able to explain additional variance in each of the two cohorts (R = 0.37 at p < 0.001 for STN; R = 0.42 at p = 0.017). Single target maps on the left correspond to renderings in figure 4 and are shown as volumetric cuts at z = −50, −30, −10, 10, 30 & 50 mm. The agreement map is shown both in volumetric and surface fashion. The BigBrain atlas served as backdrop for volumetric representations.^25^

As a final analysis, we calculated symptom-specific improvements (bradykinesia, rigidity and tremor). R-Maps calculated for rigidity and bradykinesia independently were highly similar to each other (not shown), hence they were combined to form a bradykinesia-rigidity improvement that was contrasted with tremor improvements. This goes in line with prior reports.^26^ While the R-map model for bradykinetic-rigid symptoms were similar and cross-predictive across cohorts (R = 0.48 at p < 0.01 when calculating the R-map based on STN patients to predict outcomes in the GPi cohort; R = 0.31 at p < 0.01 vice versa), they were not for tremor (STN>GPi: R = −0.28 at p = 0.11; GPi>STN: R = 0.12 at p = 0.18; figure 5). While these results could give first hints, due to the small sample size in the GPi cohort and further exclusion of 40 (STN) & 9 (GPi) patients with tremor baseline scores below two, these results should not be overinterpreted. Still, repeating the analysis with different baseline score exclusion criteria (<0-5 points) did not alter results qualitatively, suggesting that results were not dependent on this thresholding process.

**Figure 5:**
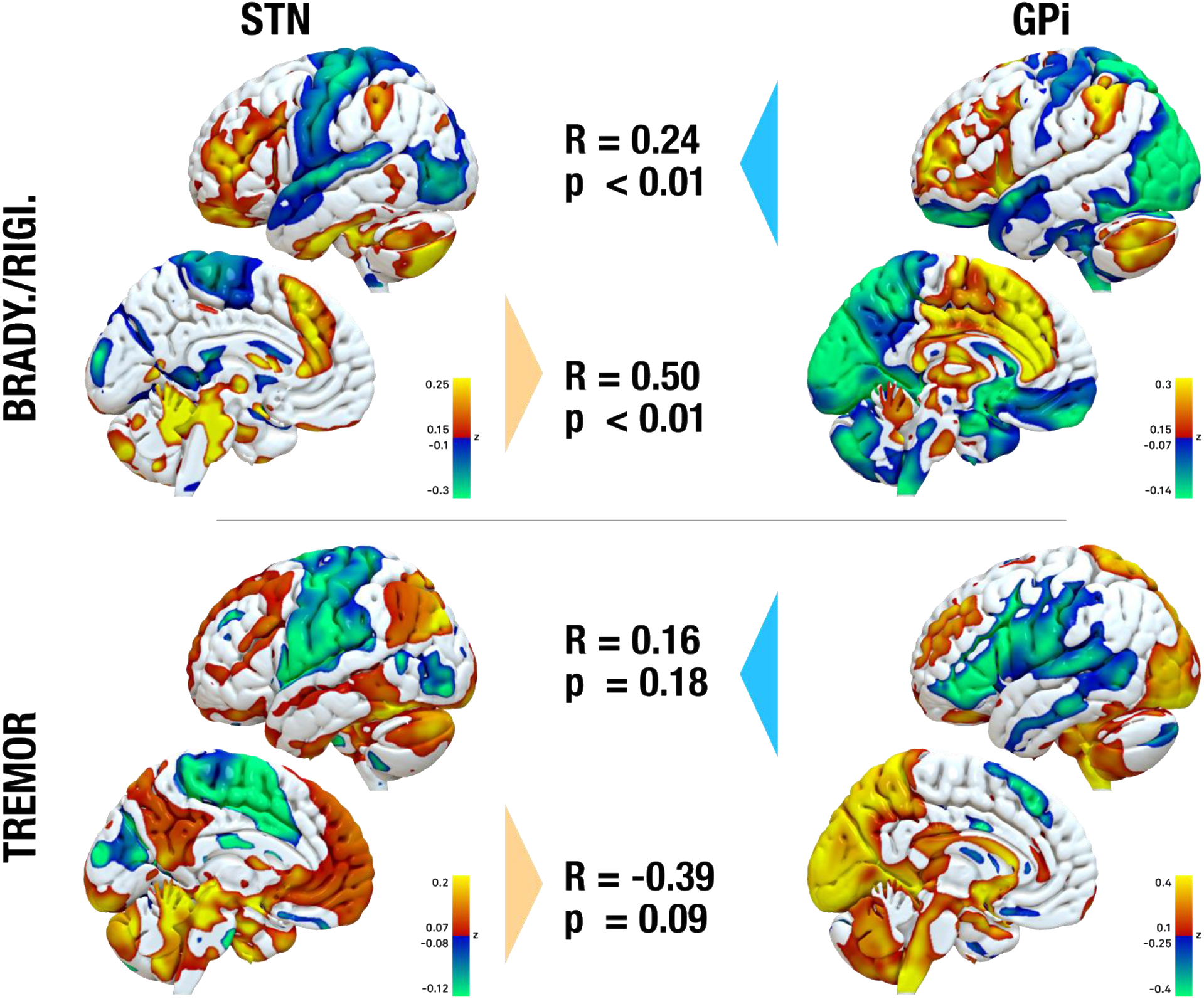
Connectivity maps associated with optimal clinical subscore outcomes. Rigidity and bradykinesia were summarized due to high similarity when analysed independently. Only this map & pattern led to significant cross-predictions across STN- and GPi-targets. Instead, effects on tremor were associated with a different connectivity pattern – also across targets and the cross-prediction did not yield significant effects.

One difference we did note across cohorts was that in the GPi cohort, especially the bradykinetic symptoms of some patients got worse under DBS. The induction of bradykinetic symptoms by GPi-DBS has been reported even in cohorts without Parkinson’s disease (such as dystonia), before.^27,28^ Hence, as an opportunity to study this relationship further, we contrasted functional connectivity profiles calculated from the six GPi patients in which bradykinesia improved most strongly (by 67.9 ±11.6 percent) with the six ones in which bradykinesia worsened most strongly (by −58.4 ±19.7 percent). This revealed significant differences in connectivity profiles. Namely, stimulation sites in worsening patients were less connected to left superior, middle and inferior frontal gyrus and more strongly connected to bilateral cerebellar and occipital regions as well as to the parieto-occipital sulcus (figure 6).

**Figure 6:**
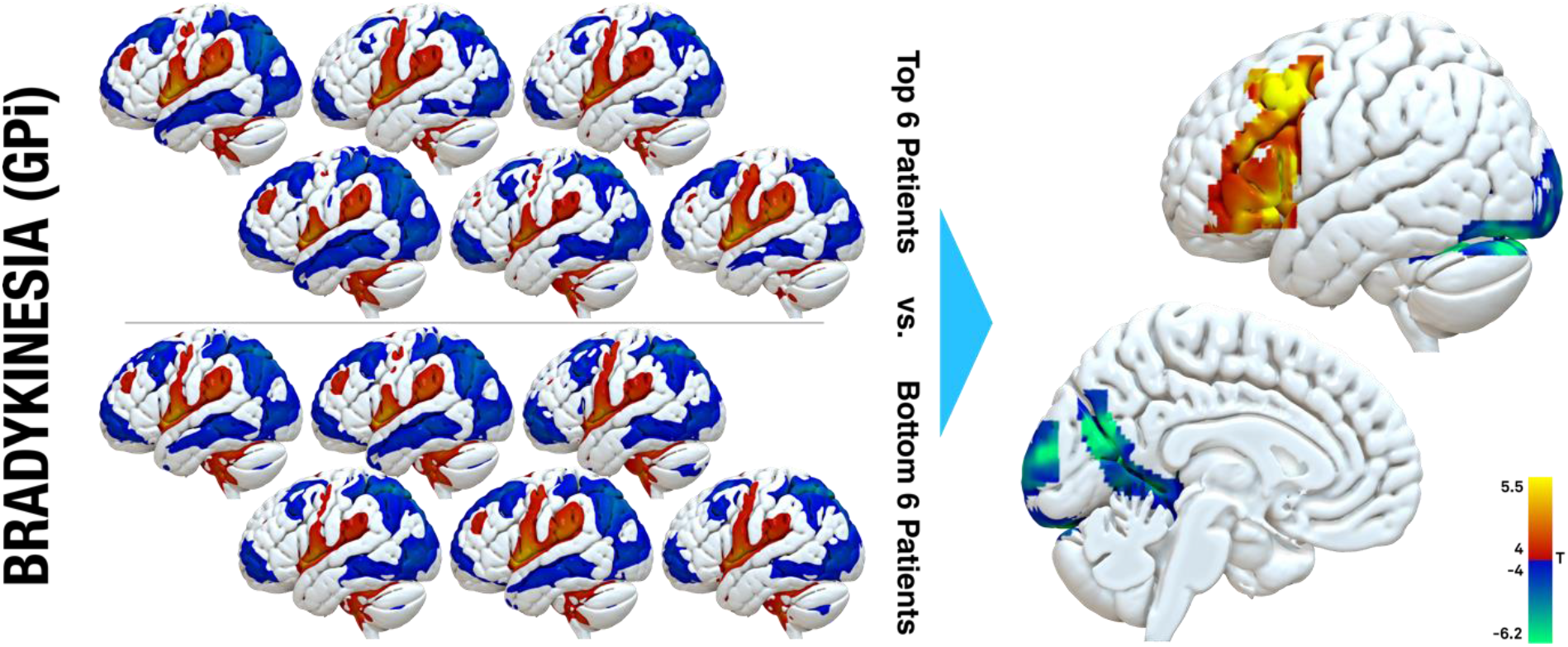
Effects of GPi-DBS on bradykinesia. Within the GPi cohort, it was noted that in some patients, bradykinetic symptoms worsened under DBS – as stated by previous reports, as well. A paired t-test contrast between connectivity fingerprints of top six improving and bottom six worsening patients revealed positive association with left middle and inferior frontal gyrus and negative association with bilateral cerebellar & occipital regions, as well as the parieto-occipital sulcus (family wise estimate corrected on cluster level at p < 0.05).

## Discussion

Three main conclusions may be drawn from this study. First, based on our results, optimal connectivity profiles of DBS to the STN and GPi for effective treatment of Parkinson’s disease are highly similar. For both targets, a functional connectivity profile with anticorrelations to sensorimotor cortices and positive connectivity to specific regions in the frontal cortex and motor cerebellum (among other regions) were associated with optimal clinical outcomes. This finding was quantified by cross-predicting across DBS targets. In other words, a model of optimal connectivity was calculated based on one target and used to predict outcomes in patients from the second target, and vice versa. This finding suggests that regardless of STN or GPi stimulation, a shared functional network could be responsible for motor symptom improvements in Parkinson’s disease. Second, we introduced a novel method to combine models of optimal network response across DBS targets (in form of an agreement map) and showed that it was able to explain variance in outcomes regardless of DBS target choice. Finally, third, our data suggests that while optimal connectivity profiles of electrodes for maximal improvements of bradykinetic-rigid symptoms were similar across targets, the ones for tremor seemed to differ.

Explanations of the pathophysiology of basal ganglia disorders such as Parkinson’s disease have been dominated by the Albin et al. and DeLong model and its subsequent modifications.^29,30^ This model describes a fronto-basal-ganglia-thalamo-cortical feedback loop, and one general concept is that inhibition of either STN or GPi due to DBS may lead to a net shift of balance leading to disinhibition of the thalamus and a propagation of motor output. In this way, effects of DBS on hypokinetic symptoms such as bradykinesia and rigidity have been explained. While this model has proven valid and powerful in many occasions, the so-called paradox of functional neurosurgery consists in the fact that both STN-DBS and GPi-DBS have equally been effective targets for hyperkinetic movement disorders such as dystonia.^31-34^

Hence, the working mechanism of the functional network targeted by DBS is thought to be more complex. A popular concept is that DBS reduces a noisy feedback-signal that may be present across the whole basal ganglia cortical loop.^31,35-37^ Namely, pathologically synchronized activity can be recorded from several sites of the basal ganglia in the beta frequency range (13-30 Hz) and is found to be supressed by either DBS or dopaminergic medication and to relate to bradykinetic-rigid symptoms (and not tremor).^38-42^ In this framework, the main mechanism of action of DBS seems to be to disrupt (pathological) information flow – likely in a specific frequency domain – throughout the motor loop.^32^ In turn, *physiological* information flow could be restored by reopening the necessary bandwidth. This could explain why both lesions and DBS to both STN and GPi are highly effective (and for both Parkinson’s disease and dystonia): A pathological noise carrier signal needs to be present for DBS to have a profound effect in that it disrupts largely monotonic and synchronized oscillating brain network activity (in case of dystonia, a similar noise signal has been described as pathologically synchronized theta activity.^43^

The method we apply here (resting-state functional MRI) is several orders of magnitude too crude to resolve aforementioned temporal dynamics. However, the present study may still contribute to this framework in form of a better *spatial* characterization of exactly *which connections* seem to matter most. Which are the underlying anatomical regions that seem to play the most crucial part in this oscillating noise loop.

Not only are the optimal network maps calculated from the two DBS targets highly similar (figure 4), but there are even specific brain regions that are quantifiably predictive of clinical outcome when modulating either of the two targets (described as the agreement map). The agreement map includes connections that i) exist in both the STN and the GPi cohort and ii) are associated with a good clinical outcome in both. It hence constitutes a model of areas to which the DBS electrode should be connected to maximize its treatment capability – regardless of the DBS target. When seen from the angle of a noisy feedback model described above, we could speculate that this circuitry might be exactly where the noisy signal should be most predominantly expressed. A gap that makes it close to impossible to directly relate connectivity information derived from resting-state fMRI and electrophysiological recordings in the beta range, however, is time. As mentioned above, temporal resolution differs by orders of magnitude and make the two signals measured by either method a completely different one. Still, some indirect hints shows that the networks identified with two methods may converge. First, we know that optimal stimulation in the STN is reached within its sensorimotor functional zone which is where beta power and STN-cortical beta coherence at rest is strongest.^16,26,44-49^ Second, regions with maximal expression of beta power were most strongly connected to prefrontal regions in a diffusion MRI based tractography study that compared connectivity profiles seeding from contacts with high beta activity to others.^50^

Accumulating evidence suggests that while the effective network responses for maximal treatment of bradykinesia and rigidity are overlapping, the ones for tremor are more distinct.^26,44^ Here, some studies attribute the effect on tremor to structures outside the STN such as the dentatorubrothalamic tract.^51,52^ In contrast, the main effect on bradykinetic-rigid symptoms has been located to a highly agreeing region within the dorsolateral STN proper by multiple groups, world-wide.^16,44,53-55^ In line with this segregation of symptoms, elevated beta power in the STN was found to correlate with severity of bradykinetic-rigid symptoms but not tremor.^56^

The divergence in responsive connectivity we observe in the current data is consistent with the existence of akinetic/rigid and tremor dominant subtypes of Parkinson’s disease. While the pathogenesis of the first type can be described using the conventional theory of an amplified balance in favor of the indirect pathway (at cost of the direct pathway), tremor is attributed a compensatory cause downwards of the stream, involving a cerebello-thalamic network.^57-59^ Our current set of datapoints is unable to characterize a tremor-network in Parkinson’s disease by good evidence. What we note is that networks calculated for bradykinesia seem similar across targets (and can be used to cross-predict outcomes) while the ones for tremor are not. Augmenting such an analysis with additional cohorts could one day lead to a personalized DBS targeting the network tailored to the leading symptom in each patient.^6^

Finally, previous studies have reported for GPi-DBS in dystonia to in fact *induce* bradykinetic symptoms.^27,28^ In direct comparison with the two cohorts shown here, we noted that bradykinetic symptoms became worse in a larger fraction of patients from the GPi-DBS cohort. When contrasting DBS connectivity profiles in the six patients in which bradykinetic symptoms worsened most with the ones in which it improved most, connectivity to cerebellar, occipital and parieto-occipital regions was associated with symptom worsening. The parieto-occipital sulcus specifically has been reported to be involved in finger-tapping tasks in patients suffering Parkinson’s disease, especially when contrasting externally vs. self-paced movements.^60^ Similar to findings relating to subscores, these explorative findings should not be overinterpreted since the study was not designed to exactly address this question. Instead, both could be helpful to form hypotheses that could be addressed in future work.

Several limitations apply to this study. First and foremost, the two cohorts were not balanced in size. Since the results of the STN cohort were used from a larger prior publication that the present paper builds upon, this could not easily be accounted for. Despite the predominant agreement of the R-maps, there are regions that remain incongruent. Especially connectivity to medial frontal gyrus and paracentral lobulus differed fundamentally between the two targets. This might be explained by brain areas that are not part of a causal network, but that were spuriously connected to DBS electrodes that led to high rates of response. Since the spontaneous activity of all connectomic voxels is recorded during a resting state functional MRI, all regions of the brain will either be functionally correlated or anticorrelated with the electrodes to some degree. Rs-fMRI itself constitutes an indirect measure of slow-dynamic brain connectivity.

Relationships to causal effects of DBS should not be drawn thoughtlessly (and if not supported by additional means of data, also see discussion above). Along the same lines, we applied *normative* connectome atlases to estimate connectivity. These datasets do not account for individual variations of connectivity but rather ask the question which connectivity profile an electrode at a specific site would have *in an average* human brain. This is a crucial distinction that should not be forgotten and largely impacts interpretation of our results. Still, use of normative connectomes in context of DBS has led to models that were predictive in out-of-sample datasets.^5,6,8,61-64^ Finally, electrode models themselves have inaccuracies that are based on factors such as i) resolution constraints of underlying data, ii) complexity of and small size of subcortical targets and iii) registration errors when aggregating data across patients to make them comparable.^16^ To minimize impact of these constraints, we used a specialized DBS imaging pipeline that is based on multispectral normalizations evaluated for both STN and GPi, phantom-validated electrode localizations and careful manual inspection of results in each processing step.^18,19,65^ The electric field model around the electrode has limitations especially when modelling bipolar stimulation settings which were only applied in 6 out of 122 patients in the current dataset.^66^ Subsequent studies should aim at confirming results when applying more advanced electrode modelling concepts such as path activation or driving force models.^67-69^

## Conclusions

In summary, our results illustrate a candidate network that could be responsible or at least associated with optimal clinical improvements in DBS for Parkinson’s disease – regardless of target choice. By pinpointing the network associated with optimal outcomes from two different nodes (STN and GPi), we were likely able to identify regions that have higher probability to be causally involved. As such, a joint (agreement) model informed by both DBS targets was able to explain larger amounts of variance in clinical outcomes in patients operated with either target. Upon further validation, this set of brain regions could potentially inform neuromodulation targets associated with clinical improvements in Parkinson’s disease above and beyond the field DBS.

## Abbreviations

STN: subthalamic nucleus
GPi: internal globus pallidus
DBS: deep brain stimulation

## Acknowledgements

Data were provided [in part] by the Brain Genomics Superstruct Project of Harvard University and the Massachusetts General Hospital, (Principal Investigators: Randy Buckner, Joshua Roffman, and Jordan Smoller), with support from the Center for Brain Science Neuroinformatics Research Group, the Athinoula A. Martinos Center for Biomedical Imaging, and the Center for Human Genetic Research. 20 individual investigators at Harvard and MGH generously contributed data to GSP Open Access Data.

## STROBE statement: Reporting guidelines checklist for cohort, case-control and cross-sectional studies

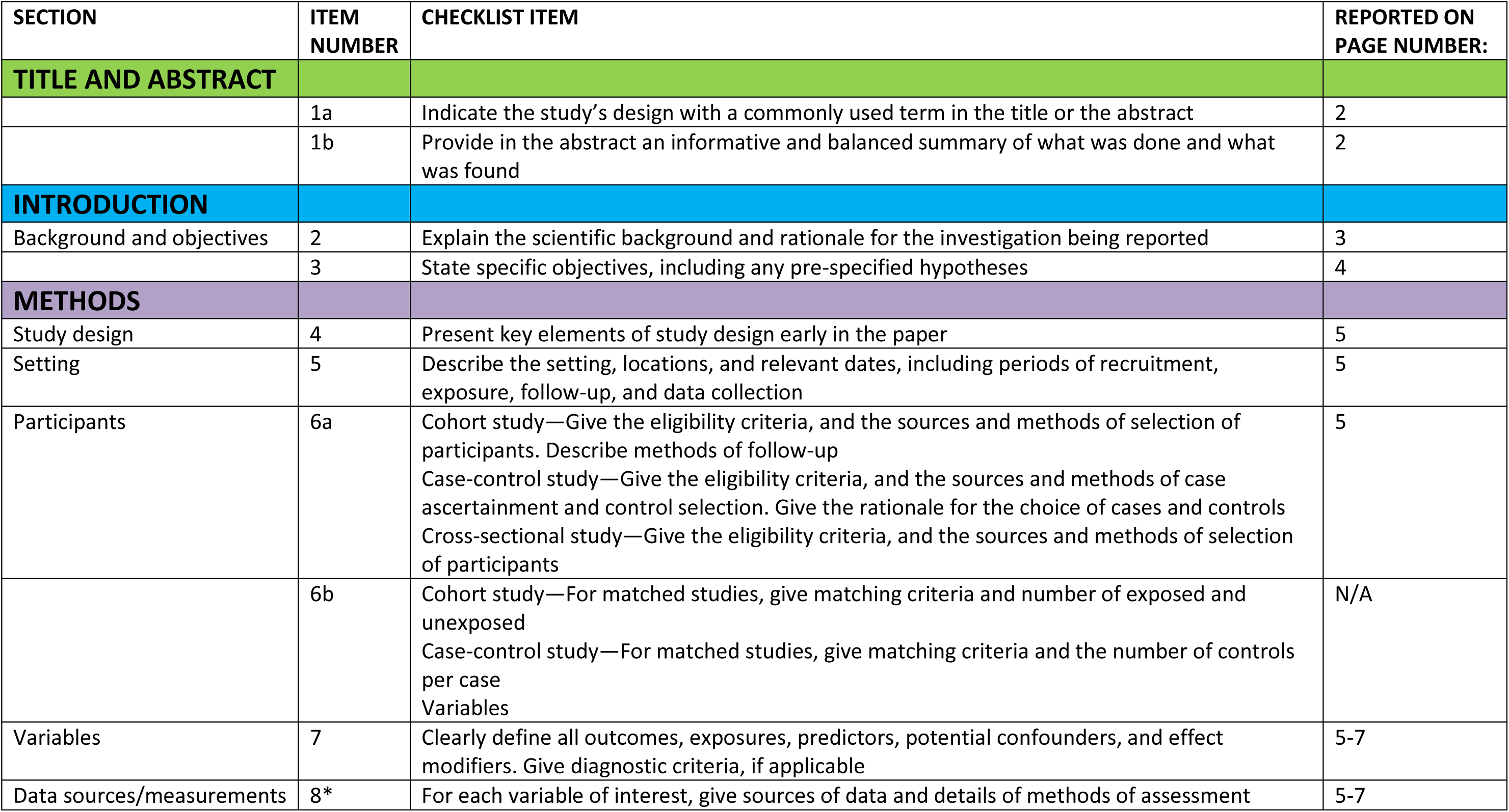
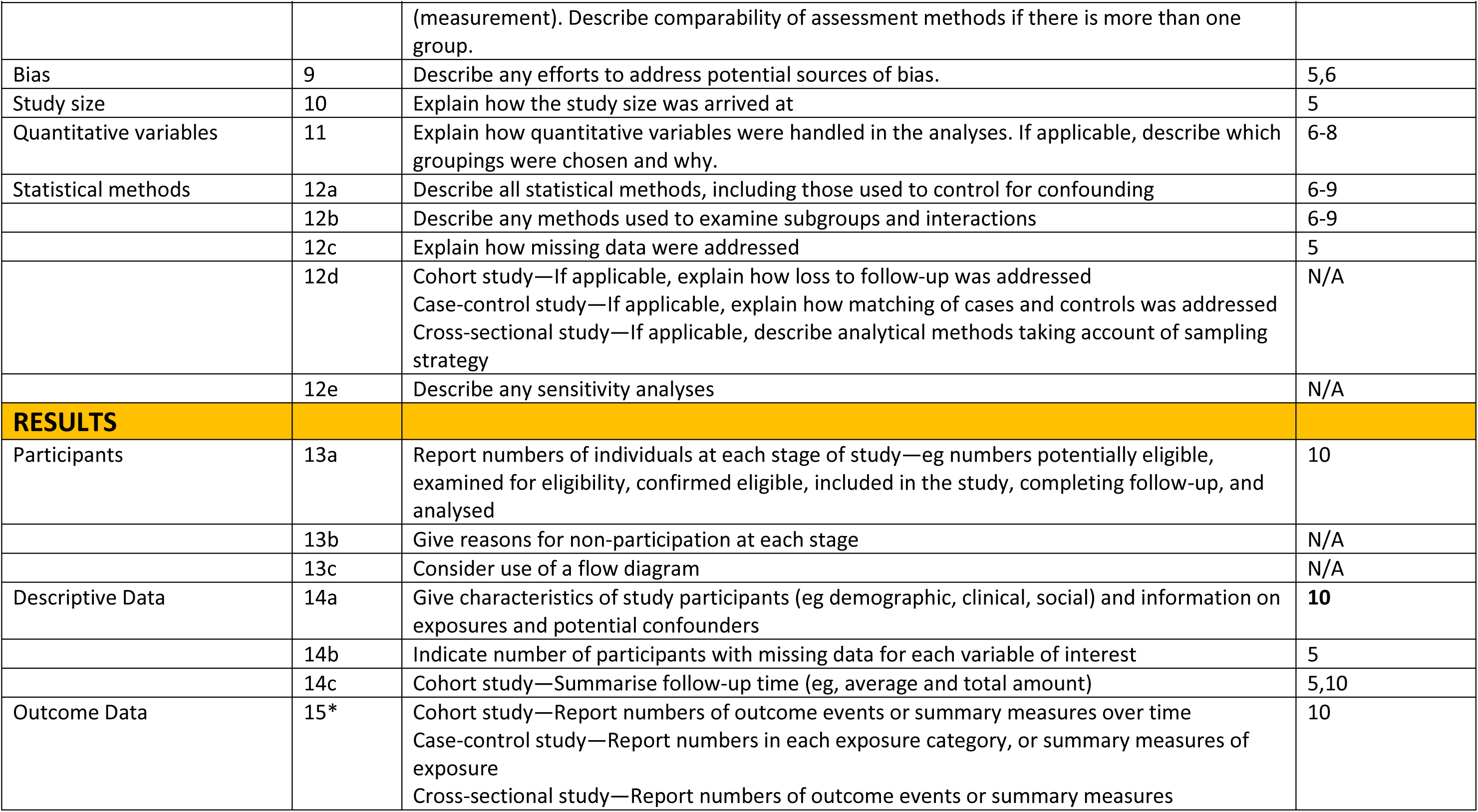
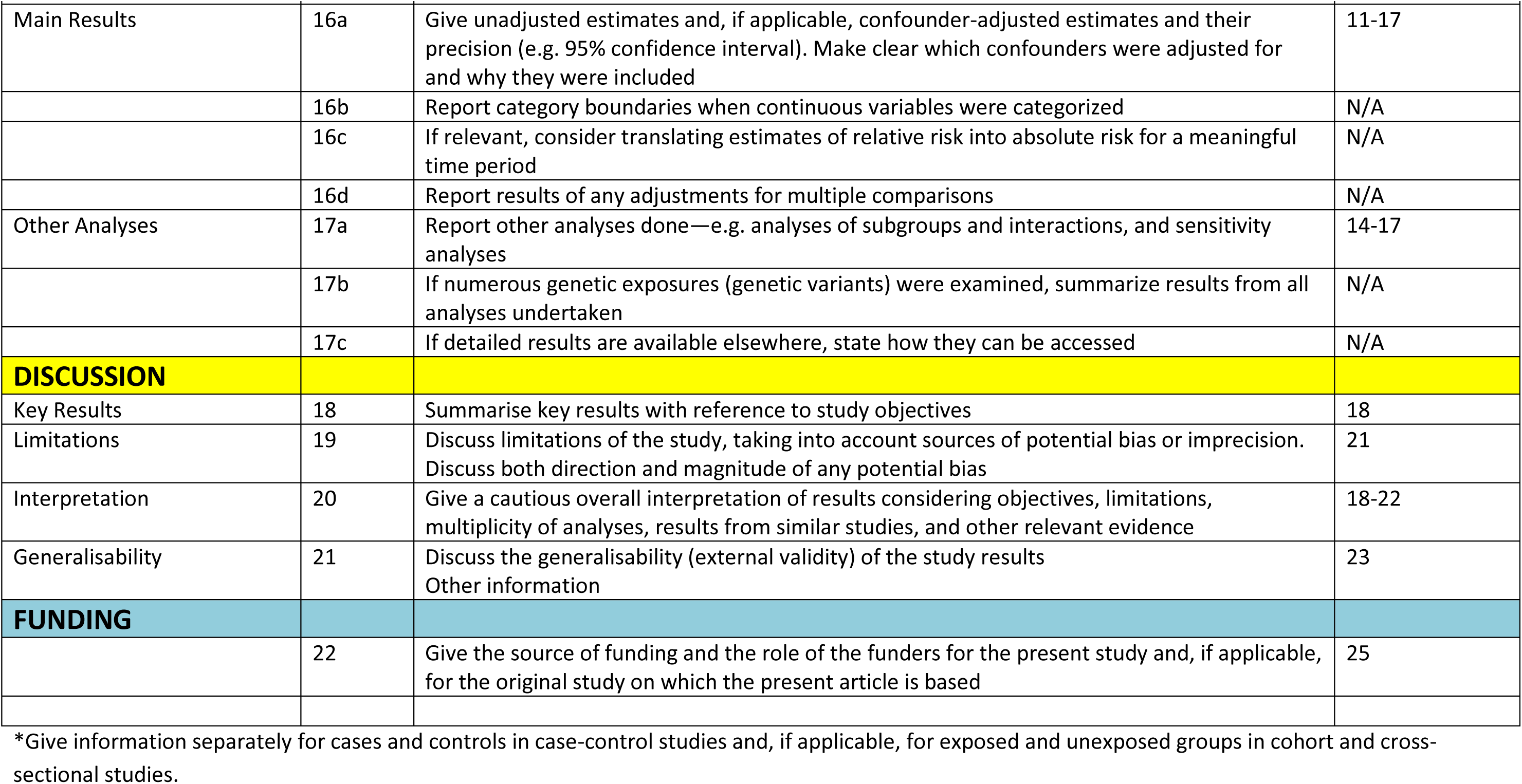

